# Mediation of the relationship between air pollution and dementia: A UK Biobank study

**DOI:** 10.64898/2026.02.20.26346698

**Authors:** Katherine Taylor, Malcolm Harris, Esther Hui, Emma Anderson, Naaheed Mukadam

## Abstract

**Background:** Air pollution is a potentially modifiable risk factor for dementia with a population attributable risk fraction of 3%. Little is known about the causal mechanisms behind the association, so we aimed to investigate this.

**Methods:** Data from the UK Biobank were used to investigate the association between six measures of air pollution (NO_2_, NO_x_, PM_2·5-10_, PM_2·5_, PM_2·5 absorbance_ and PM_10_) and dementia incidence. Indirect pathways through four mediators (cardiovascular conditions, mental health treatment, insufficient exercise and social isolation) were explored. Logistic regression was used to model the associations between air pollution, mediators and dementia. Casual mediation analysis implemented using the g-formula was used to investigate the joint indirect effect through the mediators.

**Findings:** Exposure to the highest quintile of PM_2·5_ (Rte:1·14, 95% CI:1·06-1·23), NO_x_ (Rte:1·11, 95% CI:1·03-1·20) or NO_2_ (Rte:1·08, 95% CI:0·99-1·16), compared to the lowest quintile, was associated with higher dementia risk. Most of the observed association resulted from the direct effect of air pollution, consisting of pathways not captured through considered mediators. Amongst those in the highest PM_2·5_ quintile, jointly intervening on the four mediators would result in a 1% reduction in risk of dementia (Rpnie:1·01, 95% CI: 1·01-1·02). The randomised pure natural indirect effect was similar for NO_2_ (Rpnie:1·01, 95% CI: 1·00-1·01) and NO_x_ (Rpnie:1·01, 95% CI: 1·01-1·02).

**Interpretation:** Most of the association between dementia and PM_2·5_, NO_2_ and NO_x_ occurs through the direct effect of air pollution, or other unmeasured mediators, and not pathways through these four mediators.

**Funding:** Medical Research Council (Grant MR/W006774/1).

## Introduction

There were an estimated 982,000 older people living with dementia in the UK in 2024, costing the UK an estimated £42 billion per annum(1). Therefore, identification of points for intervention to attenuate this population health challenge is vital.

Air pollution is a significant public health concern, with higher levels being associated with negative health outcomes(2). There is growing evidence supporting an association between higher levels of air pollution and increased incidence of dementia in older adults(3,4). The 2024 Lancet Commission on Dementia Prevention, Intervention and Care listed air pollution as one of 14 potentially modifiable risk factors associated with dementia, calculating its population attributable risk fraction to be 3% (5).

The causal mechanisms behind the association between air pollution and dementia are still undetermined. One hypothesis suggests that ultrafine particles pass through the blood-brain barrier via the fluid of the lungs and the respiratory tract(6). Once particulate matter has reached the brain, oxidative stress caused by the generation of free radicals may result in neurodegeneration(7). Alternatively, identification of mediators on the causal pathway from air pollution to dementia could provide an insight into casual mechanisms. It would also highlight potential points for intervention, enabling us to attenuate the association between air pollution and dementia.

There has been limited research into mediation of this pathway. Given the wide-ranging health impacts of air pollution and the multiple potentially modifiable risk factors for dementia, it is possible that indirect pathways through health and lifestyle factors exist. Existing literature has considered the potential for cardiovascular health to mediate the association, with one study finding evidence for partial mediation through cardiovascular disease. They observed cardiovascular disease to mediate approximately 9% and 21% of the association between dementia, NO_2_ and PM_2·5_ respectively(8). Another study using a Swedish cohort found that only a small proportion of the association between PM_2·5_ and dementia was through a direct effect after taking into account indirect effects through cardiovascular health. However, this study did not use a counterfactual framework as recommended when dealing with a binary outcome(9).

Existing literature focuses on one mediator at a time however, given that multiple health and social factors are associated with both air pollution and dementia, it is likely that multiple mediatory pathways exist(2,5). The present study aims to address gaps in the literature by exploring the indirect pathways from air pollution to dementia through multiple mediators. We used the UK Biobank to investigate associations between dementia incidence and six measures of air pollution. Following this we considered the potential for cardiovascular health, exercise, social interaction, and mental health to mediate these associations. Our analysis provides insight into the causal mechanisms linking air pollution and dementia and highlights routes for intervention which could result in a reduced incidence of dementia among those exposed to high levels of air pollution.

## Methods

### Sample selection

The UK Biobank (UKB) is a large-scale prospective study that recruited 502,180 participants aged 40-69 between 2006-2010 (10). Details regarding the sample and collection methods have been described elsewhere (10). UKB received ethical approval from the Northwest Multi-Centre Research Ethics Committee (11/NW/0382 and 16/NW/0274). Data were analysed under UKB application number 123335.

Complete case analysis was used. Participants with dementia prior to the measurement of air pollution data were excluded from this analysis. Participants living more than 400km from Greater London were not included due to concerns about data reliability for particulate matter measures.

### Exposure

Six measures of residential air pollution were obtained: NO_2_, NO_x_, PM_10,_ PM_2·5-10_, PM_2·5_, (µg/m^3^) and PM_2·5 absorbance_ (parts per metre). Measures were based on participants’ home postcode at baseline. All residential air pollution measures were modelled based on a Land Use Regression model, based on monitoring conducted between January 2010 and January 2011(11). Quintiles of each air-pollution measure were generated and used as the exposure within this analysis.

### Outcome

Our outcome was all-cause dementia. Diagnosis was obtained through algorithmic combination of self-reported data at baseline assessment, hospital admission diagnoses, and ICD-10 codes in death register records (12). Cases were defined using UK Biobank’s algorithmically defined diagnostic variables (13).

### Mediators

We considered four potential mediating variables: cardiovascular health, exercise, mental health, and frequency of social contact.

A measure of cardiovascular health was developed from diagnosis of six cardiovascular health conditions: angina pectoris; cardiac arrest; heart failure; essential hypertension; stroke; and diabetes. Self-reported diabetes was measured at baseline, and other cardiovascular health diagnoses were algorithmically defined based on self-reported data and linkage data from medical records(14). A binary categorisation was created identifying those with a history of any of the six cardiovascular health conditions considered.

Physical activity was measured at the baseline assessment. Participants were asked about how much moderate and vigorous activity they undertook per day. Participants were classified according to whether or not they met WHO guidelines for sufficient exercise(15).

At baseline participants were asked whether they had previously seen a psychiatrist for symptoms of depression or anxiety. Responses were categorised into a binary mental health service utilisation variable.

Frequency of social interaction was measured through frequency of friend/family visits at baseline. Responses were recorded categorically (never/almost never, once every few months, once a month, once a week, two to four times a week, almost daily) and binarized according to whether they received social visits at least once a month(16).

### Confounders

All confounders were self-reported and measured at the baseline assessment. Age and Townsend deprivation index were treated as continuous measures. The Townsend deprivation index was calculated based on a participant’s postcode, combining z-scores for the percentage of households without a car, overcrowded households, households not owner occupied and persons unemployed in a postcode. A positive score indicates material deprivation and a negative score, relative affluence (17). Sex was categorised into male and female. Ethnicity was categorised into White or Other Ethnicities. Employment status was categorised into working/studying/volunteering full or part-time and not in work for any reason. Highest educational qualification was categorised into no qualifications, GCSE or equivalent, A/AS level or equivalent and degree or other professional qualification. Smoking status was categorised into smoker and non-smoker.

### Preliminary analysis

Statistical analyses were carried out using STATA version 17 and RStudio Version 2025.09.2+418.

A series of logistic regressions were conducted to investigate the association between air pollution measures and all-cause dementia. Further analyses were restricted to air pollution exposures where associations with dementia were observed.

We expect variables which mediate the association between air pollution and dementia to be associated with both air pollution and dementia. For each mediator, we ran a logistic regression model observing the association between air pollution and the mediator and a second logistic regression model to observe the association between the mediator and dementia. All analyses were adjusted for all confounders as previously described.

### Mediation analysis

The proposed causal pathway from air pollution measure to dementia is shown in Figure 1. Within our analysis all pathways were modelled using logistic regression and adjusted for confounders as previously described. The CMAverse package was used to conduct a causal mediation analysis. G-formula was selected as the modelling approach. The estimation procedure has been described elsewhere(18–20). Briefly, this approach works by considering the impact of jointly intervening on the distribution of mediators rather than simply setting the mediator value for each participant to be that expected in a world where everyone had been exposed to the lowest level of air pollution. Indirect effects can be interpreted as the impact of jointly intervening on the considered mediators. 95% CI were generated using 1000 bootstrap samples. E-values were calculated to evaluate the sensitivity of decomposed effects to unmeasured confounding(21).

**Figure 1.**
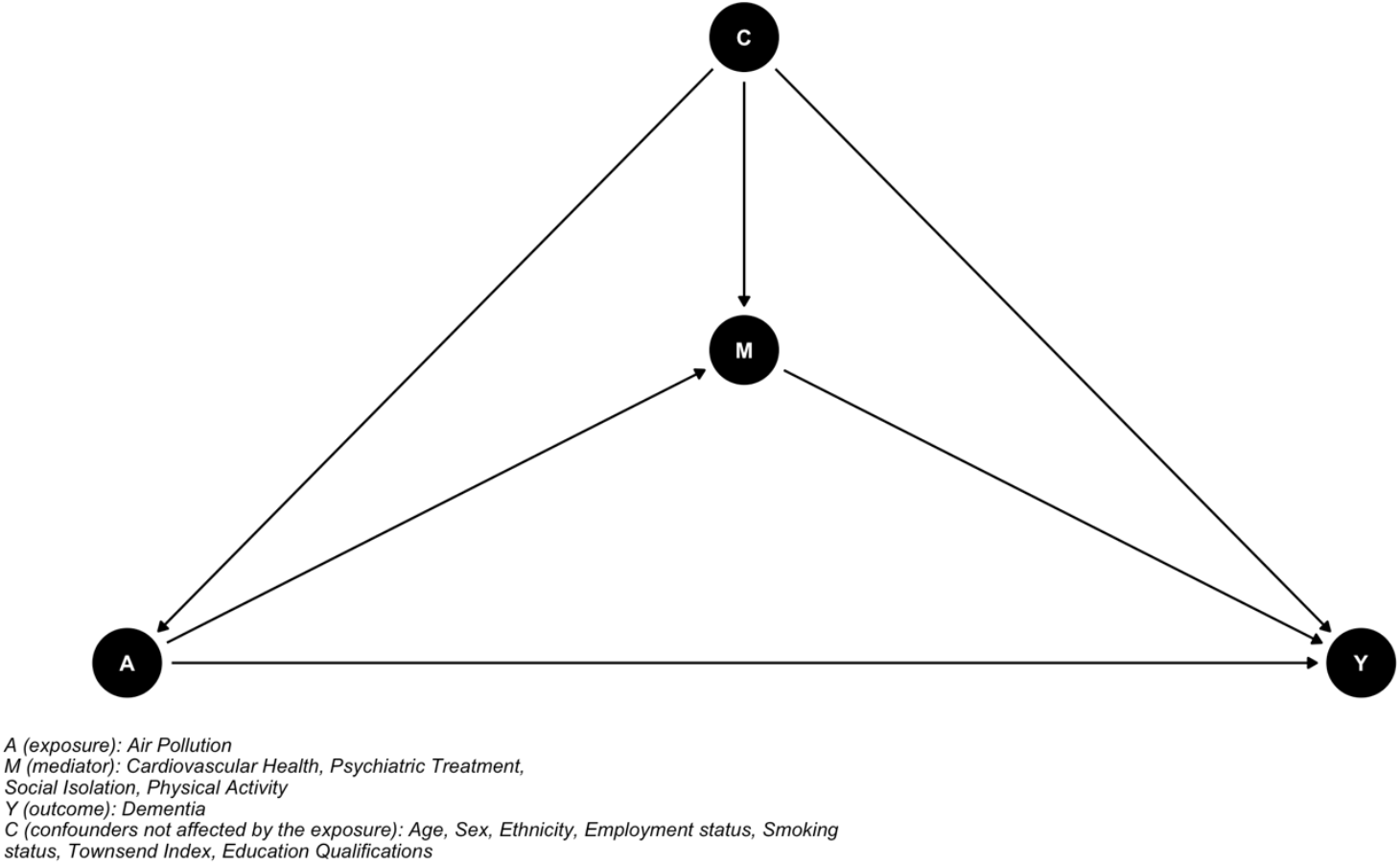
Directional Acyclic Graph of the causal pathway from air pollution to dementia through cardiovascular and mental health.

## Results

Following exclusion of participants who did not meet our inclusion criteria, our analytical sample consisted of 432,935 participants (Supplementary Figure 1). 8,137 (1·88%) participants received a dementia diagnosis during follow-up (Table 1).

**Table 1.** Sample characteristics.

| Characteristic | Category | Incident dementia<br>(N=8137) |  | No dementia<br>(N=424,798) |  |
| --- | --- | --- | --- | --- | --- |
| Demographic Characteristics |  |  |  |  |  |
| Age at baseline |  | 64·24 (4·69) |  | 56·42 (8·06) |  |
| Sex | Males | 4,227 | 51·95% | 193,332 | 45·51% |
|  | Females | 3,910 | 48·05% | 231,466 | 54·49% |
| Ethnicity | White | 7,823 | 96·14% | 402,906 | 94·85% |
|  | Other ethnicities | 314 | 3·86% | 21,892 | 5·15% |
| Qualifications | No qualifications | 2,834 | 34·83% | 69,994 | 16·48% |
|  | GCSEs/O levels or equivalent | 1,823 | 22·40% | 118,280 | 27·84% |
|  | AS/A levels or equivalent | 1,279 | 15·72% | 76,283 | 17·96% |
|  | Degree/professional qualifications | 2,201 | 27·05% | 160,241 | 37·72% |
| Employment Status | Full/part-time paid or unpaid work/student | 1,987 | 24·40% | 266,799 | 62·80% |
|  | Not in work | 6,150 | 75·60% | 157,999 | 37·20% |
| Smoking Status | Non-smoker | 7324 | 90·00% | 381,285 | 89·8% |
|  | Smoker | 813 | 10·00% | 43,513 | 10·2% |
| Townsend Deprivation Index |  | -1·09 (3·20) |  | -1·40 (3·01) |  |
| Air Pollution Characteristics – N and % |  |  |  |  |  |
| NO<sub>2</sub> (µg/m<sup>3</sup>) | Quintile 0 | 1,451 | 17·8% | 85,146 | 20·0% |
|  | Quintile 1 | 1,647 | 20·2% | 84,976 | 20·0% |
|  | Quintile 2 | 1,776 | 21·8% | 84,933 | 20·0% |
|  | Quintile 3 | 1,659 | 20·4% | 84,890 | 20·0% |
|  | Quintile 4 | 1,604 | 19·7% | 84,853 | 20·0% |
| NO<sub>x</sub> (µg/m<sup>3</sup>) | Quintile 0 | 1,457 | 17·9% | 85,168 | 20·0% |
|  | Quintile 1 | 1,662 | 20·4% | 85,168 | 20·0% |
|  | Quintile 2 | 1,647 | 20·2% | 84,950 | 20·0% |
|  | Quintile 3 | 1,710 | 21·0% | 84,807 | 20·0% |
|  | Quintile 4 | 1,661 | 20·4% | 84,892 | 20·0% |
| PM2·5 (µg/m<sup>3</sup>) | Quintile 0 | 1,459 | 17·9% | 86,243 | 20·3% |
|  | Quintile 1 | 1,631 | 20·0% | 84,888 | 20·0% |
|  | Quintile 2 | 1,636 | 20·1% | 85,191 | 20·1% |
|  | Quintile 3 | 1,721 | 21·2% | 83,741 | 19·7% |
|  | Quintile 4 | 1,690 | 20·8% | 84,735 | 19·9% |
| PM10 (µg/m<sup>3</sup>) | Quintile 0 | 1,489 | 18·3% | 85,479 | 20·1% |
|  | Quintile 1 | 1,703 | 20·9% | 86,011 | 20·2% |
|  | Quintile 2 | 1,709 | 21·0% | 83,512 | 19·7% |
|  | Quintile 3 | 1,661 | 20·4% | 84,912 | 20·0% |
|  | Quintile 4 | 1,575 | 19.4% | 84,884 | 20.0% |
| **PM2.5-10 ($\mu\text{g}/\text{m}^3$)** | Quintile 0 | 1,596 | 19.6% | 85,174 | 20.1% |
|  | Quintile 1 | 1,713 | 21.1% | 87,704 | 20.6% |
|  | Quintile 2 | 1,625 | 20.0% | 83,458 | 19.6% |
|  | Quintile 3 | 1,623 | 19.9% | 84,161 | 19.8% |
|  | Quintile 4 | 1,623 | 19.4% | 84,301 | 19.8% |
| **PM2.5 absorbance ($\text{m}^{-1}$)** | Quintile 0 | 1,609 | 19.8% | 90,638 | 21.3% |
|  | Quintile 1 | 1,642 | 20.2% | 82,982 | 19.5% |
|  | Quintile 2 | 1,816 | 22.3% | 86,007 | 20.2% |
|  | Quintile 3 | 1,571 | 19.3% | 83,208 | 19.6% |
|  | Quintile 4 | 1,499 | 18.4% | 81,963 | 19.3% |
| **Mediator Characteristics** |  |  |  |  |  |
| **Cardiovascular conditions** | 0 | 2,287 | 28.11% | 239,970 | 56.49% |
| | $\geq 1$ | 5,850 | 71.89% | 184,828 | 43.51% |
| **Meeting WHO exercise guidelines** | Yes | 7,566 | 92.98% | 394,874 | 92.96% |
|  | No | 571 | 7.02% | 29,924 | 7.04% |
| **Mental health service utilisation** | Yes | 6,907 | 15.12% | 47,741 | 11.24% |
|  | No | 1,230 | 84.88% | 377,057 | 88.76% |
| **Family/friend visits** | Less than once per month/never | 755 | 9.28% | 35,246 | 8.30% |
|  | At least once per month | 7,382 | 90.72% | 389,552 | 91.70% |

Participants with dementia were more likely to be older, male and White. Quintiles of air-pollution did not differ by dementia status. Participants with dementia were more likely to have a cardiovascular condition (71·89% compared to 43·51%) and more likely to have seen a psychiatrist for symptoms of anxiety or depression (15·12% vs 11·24%) (Table 1).

We observed that higher levels of PM_2·5_ were associated with increased odds of dementia. Individuals in the highest quintile of PM_2·5_ exposure had 15% higher odds of developing dementia compared to those in the lowest quintile (OR:1·15 95% CI:1·06-1·24). Higher levels of NO_x_ were also associated with a higher incidence of dementia. Individuals who were exposed to the highest quintile of NO_x_ had 8% increased odds of developing dementia compared to those in the lowest quintile (OR: 1·12, 95% CI: 1·04-1·21). Higher levels of NO_2_ were associated with higher odds of developing dementia. We do not observe a dose-response relationship between dementia and PM_2·5,_ NO_x_ or NO_2_. The OR of developing dementia in those exposed to higher than the lowest quintile of air-pollution was similar in quintiles one to four, with the 95% CI overlapping between the exposure categories (Figure 2, Supplementary Table 1).

**Figure 2.**
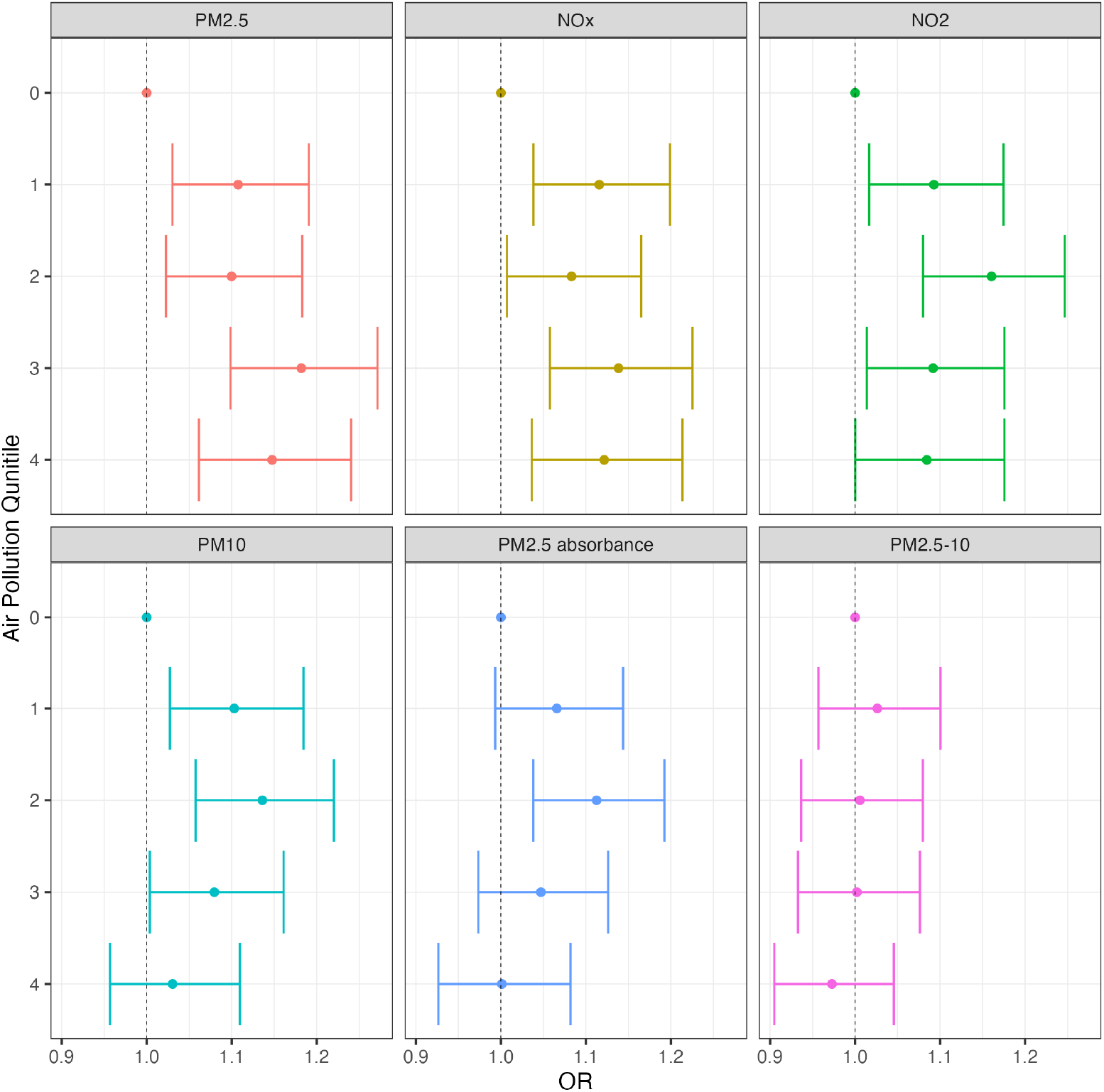
Odds ratio of dementia incidence in association with air pollution quintile.

We found some evidence for an association between dementia, PM_10_ and PM_2·5 absorbance_. Individuals in PM_10_ quintiles one and two had significantly higher odds of developing dementia compared to those in the lowest air pollution quintile (Q1 – OR:1·10, 95% CI: 1·03-1·18. Q2 – OR: 1·14, 95% CI: 1·06-1·22). Individuals in PM_2·5 absorbance_ quintile three had higher odds of developing dementia compared to those in the lowest air pollution quintile (OR:1·11, 95% CI:1·04-1·19). However, there was no difference in odds of dementia in those exposed to the highest levels of PM_10_ and PM_2·5 absorbance_ compared to those in the lowest quintile. We found no evidence of a relationship between dementia incidence and PM_2·5-10_ and so we excluded this exposure from further analyses (Figure 2, Supplementary Table 1).

Each of the mediators considered within this study was associated with higher odds of dementia incidence (Supplementary Figure 2). Those with at least one cardiovascular condition had 71% higher odds of developing dementia compared to those who did not have a cardiovascular condition (OR: 1·71, 95% CI: 1·62-1·80). Having received psychiatric treatment for depression or anxiety was associated with 47% increased odds of dementia incidence (OR: 1·47, 95% CI: 1·38-1·57). Social isolation was associated with 29% increased odds of developing dementia (OR: 1·29, 95% CI: 1·19-1·39) and insufficient exercise was associated with 11% increased odds of developing dementia (OR:1·01-1·21) (Supplementary Figure 2, Supplementary Table 2).

We found variable associations between air pollution measures and potential mediators (Figure 3). Compared to being exposed to the lowest quintile, being exposed to higher levels of PM_2·5_, NO_x_ and PM_10_ was associated with higher odds of having at least one cardiovascular condition (PM_2·5_ Q4 - OR: 1·10, 95% CI: 1·07-1·12. NO_x_ Q4 – OR: 1·07, 95% CI: 1·05-1·10. PM_10_ Q4 –OR: 1·04, 95% CI: 1·02-1·06). Additionally, compared to individuals in the baseline quintile, individuals exposed to quintiles one to three of NO_2_ and PM_2·5 absorbance_ had higher odds of having at least one cardiovascular condition (NO_2_ Q3 – OR: 1·10, 95% CI: 1·08-1·12. PM_2·5 absorbance_ Q3 – OR: 1·07, 95% CI: 1·05-1·10). Higher levels of PM_2·5_, NO_x_ and NO_2_ were associated with higher odds of having sought psychiatric treatment for depression or anxiety (PM_2·5_ Q4 – OR: 1·10, 95% CI: 1·07-1·14. NO_2_ Q4 – OR: 1·11, 95% CI: 1·07-1·15. NO_x_ Q4 –OR: 1·11, 95% CI: 1·07-1·14). Dose-response relationships were observed between having sought psychiatric treatment for depression or anxiety and all air pollution measures apart from PM_2·5_. Associations between air pollution and social isolation were inconsistent. Null associations were observed between social isolation and PM_2·5_ and PM_10_. Compared to individuals exposed to the lowest quintile of air pollution, individuals exposed to quintiles one to three of NO_x_, NO_2_ and PM_2·5 absorbance_ had lower odds of being socially isolated (NO_x_ Q3 – OR:0·94, 95% CI: 0·91-0·98. NO_2_ Q3 – OR: 0·90, 95% CI: 0·87-0·93. PM_2·5 absorbance_ Q3 – OR:0·89, 95% CI: 0·86-0·93). No associations were observed between insufficient exercise and air pollution measures (Figure 3, Supplementary Table 3).

**Figure 3.**
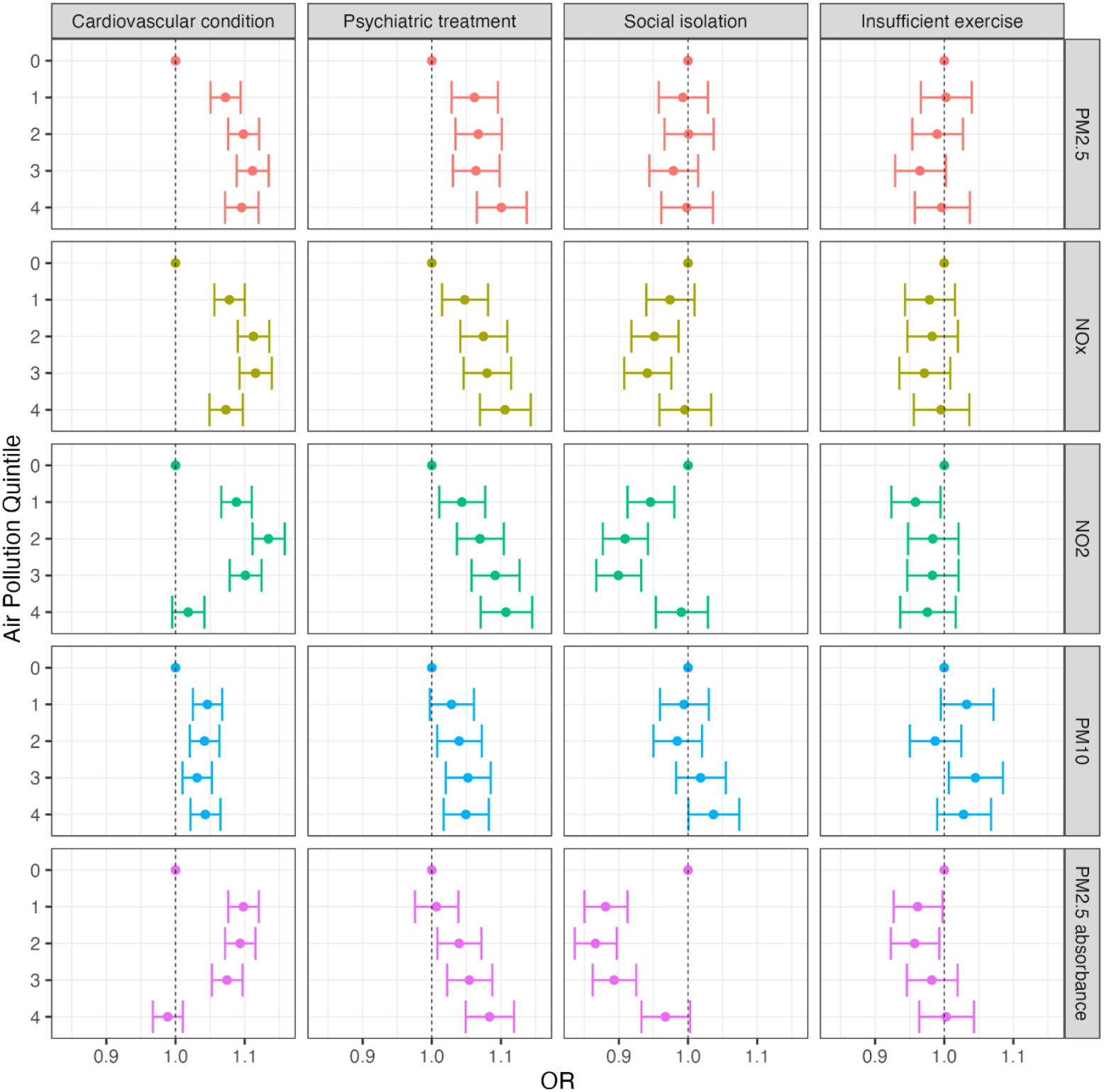
Odds ratio of mediator prevalence in association with air pollution quintiles.

Given findings from the preliminary analyses, causal mediation analysis was conducted considering NO_2_, NO_x_ and PM_2·5_ as exposures. For all measures of air pollution, being exposed to higher than the lowest quintile of air pollution was associated with a higher risk of developing dementia. Most of this association was explained by the randomised pure direct effect of air pollution on dementia. Only a small proportion of the total effect could be attributed to pathways through cardiovascular health, mental health, social isolation or physical activity (Figure 4, Supplementary Table 4).

**Figure 4.**
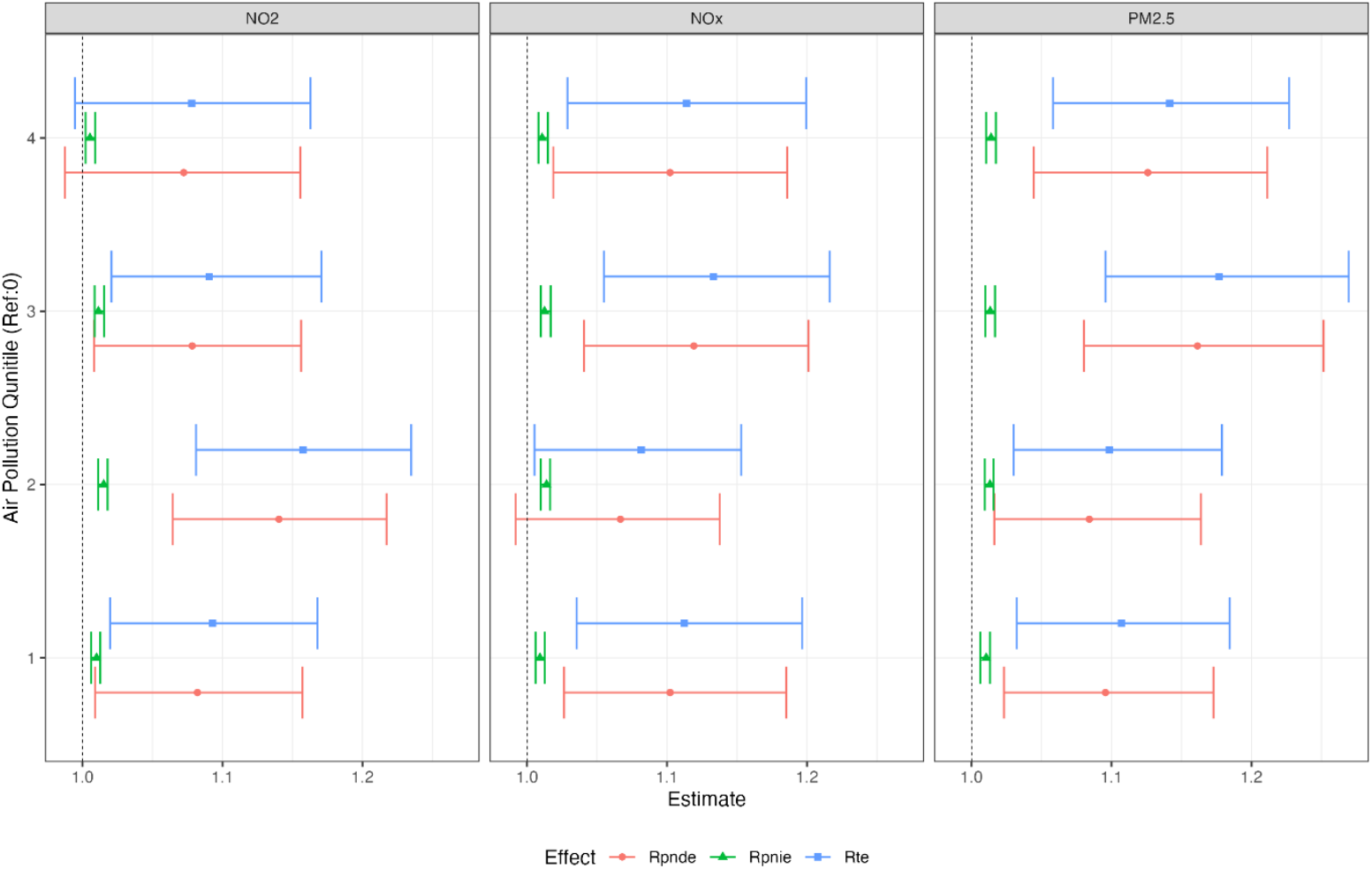
Joint mediation effects of cardiovascular health conditions and psychiatric treatment on the association between air pollution quintile and dementia. Rpnde: Randomised pure natural direct effect. Rpnie: Randomised pure natural indirect effect. Rte: Randomised total effect.

We find some evidence for mediation of the pathway from PM_2·5_ to dementia by cardiovascular health, mental health, social isolation and physical activity, though the magnitude of the randomised pure natural indirect effect (Rpnie) was small. Among those in the highest PM_2·5_ quintile, jointly intervening on mediators to shift their probability distribution to be the same as for individuals in the lowest air pollution quintile would result in a 1% reduction in risk of dementia (Rpnie:1·01, 95% CI: 1·01-1·02). The Rpnie through cardiovascular health, mental health, social isolation and physical activity was similar when NO_2_ (Rpnie:1·01, 95% CI: 1·00-1·01) and NO_x_ were considered as exposures (Rpnie:1·01, 95% CI: 1·01-1·01).

Sensitivity analyses revealed that the indirect effects observed are sensitive to unmeasured confounding. An unmeasured confounder with a relative risk of 1·11 in association with PM_2·5_ and dementia would be sufficient to explain the Rpnie estimate for the highest quintile of PM_2·5_ compared to the lowest quintile of PM_2·5_ (E value RR: 1·13, Lower 95% CI:1·11). E values for the Rpnie observed when comparing the highest and lowest quintiles of NO_x_ (E value RR: 1·11, Lower 95% CI:1·10) and NO_2_ (E value RR: 1·08, Lower 95% CI:1·05) were similar to those observed for PM_2·5_ (Supplementary Table 5).

## Discussion

### Key findings

Within UKB, compared to individuals in the lowest air pollution quintiles, those exposed to higher levels of PM_2·5_, NO_2_ and NO_x_ had higher odds of developing dementia. Most of the total effect of PM_2·5_, NO_2_ and NO_x_ on dementia risk can be attributed to pathways not captured by mediators considered in this analysis. We found some evidence for mediation by cardiovascular and mental health, physical inactivity and social isolation; however, these indirect pathways explained only a small proportion of the total effect.

### Current Literature

The total effects we observed are in line with previous literature. Existing evidence is strongest regarding the association between PM_2·5_ and dementia (22,23). A meta-analysis found that a 5 μg/m^3^ increase in exposure to PM_2·5_ was associated with an 8% increased hazard of dementia (HR: 1·08, 95% CI:1·02-1·14). Studies which considered PM_2·5_ categories also observed significant associations between higher levels of air pollution and increased dementia risk. However, only two of four studies observed a dose response relationship. A meta-analysis also revealed that a 10 μg/m^3^ increase in exposure to NO_2_ was associated with a 3% increase in hazard of dementia (HR:1·03, 95% CI 1·01–1·05). A systematic review revealed that three out of four studies which considered the relationship between categorical measures of NO_2_ and Alzheimer’s disease did not observe a dose response relationship, in line with our findings(22). Meta-analysis investigating the association between NO_x_ and dementia found that a 10 μg/m^3^ increase in NO_x_ exposure was associated with a 5% higher hazard of dementia however, the 95% CI crossed the null (95% CI 0·97–1·13) (22). We found some evidence to suggest an association between dementia and PM_10_ and PM_2·5_ absorbance, though this evidence was weaker compared to that for PM_2·5_, NO_2_ and NO_x_. Existing literature investigating these exposures is heterogeneous though meta-analysis did find a significant association between PM_2·5 absorbance_ and dementia (22).

To our knowledge, no other study has considered multiple different health and lifestyle variables as mediators on the pathway from air pollution to dementia. We find no evidence for an association between air pollution and physical activity or social isolation. Mental health was associated with our exposure and outcome so is a plausible mediator on this causal pathway. In a prior study long-term exposure to PM_2·5_, NO_2_ or NO_x_ was associated with increased risk of anxiety and depression (24). Depression and anxiety are both associated with higher levels of dementia, with depression included as a potentially modifiable risk factor in the Lancet Commission for Dementia prevention, intervention and care (5).

Most studies investigating mediation of the association between air pollution and dementia focus on PM_2·5_ (25). We found mediators only accounted for a small proportion of the association. This aligns with previous findings. Among a cohort of Canadian older adults’ cardiovascular health mediated 21% of the PM_2·5_ dementia association on a multiplicative scale and 4% an additive scale (8). Another study in a nationally representative cohort in the US found neither stroke or hypertension significantly mediated the association between dementia and PM_2·5_ (26). A Swedish study found that half the association was explained by a preceding stroke (9). However, they used a product-of-coefficients approach which is not recommended for a binary outcome such as dementia because of the non-collapsibility of odds ratios (27).

We expand on existing literature by investigating pathways to dementia from NO_2_ and NO_x_. A previous study supports our findings, showing cardiovascular health mediated 9% of the NO_2_– dementia association on a multiplicative scale and 2% on an additive scale (8). This leaves a large proportion of the association unexplained by mediation through cardiovascular health. Another study found no evidence for mediation of the association between NO_X_ and dementia by cardiovascular health (9). These studies support our findings that most of the impact of air pollution on dementia via pathways not captured through consideration of cardiovascular health, mental health, physical inactivity and social isolation.

Given we found limited evidence for mediation, mechanisms through which air pollution directly increased dementia risk should be investigated. Oxidative stress, caused by particulate matter passing through the blood-brain barrier and producing free radicals that cause neural degradation in the brain, could be the causal mechanism underpinning the air pollution-dementia association (7). The fact that PM_2·5_, defined as the concentration of all particles smaller than 2·5 microns in diameter, is strongly associated with increased dementia prevalence, whereas PM_2·5-10_, which measures particles between 2·5 and 10 microns in diameter, does not have this association, lends further support to this theory as bigger particles may be less able to cross the blood brain barrier. Additionally, in mediation analyses neuroinflammation, represented by cerebrospinal fluid triggering receptor expressed on myeloid cell two (CSF TREM2), was observed to partially mediate the association between PM_2·5_ exposure and aggravated deposition of amyloid (28). Differential DNA methylation related to neuroinflammation was also found to mediate the association between traffic‐related PM_2·5_ and Alzheimer’s disease (29).

### Strengths and limitations

This paper considers a range of potentially modifiable mediators on the causal pathway from air pollution to dementia. Additionally, we were able to consider six different measures of air pollution and consider the potential for mediatory pathways to differ dependent on the exposure. Our choice of mediation analysis methodology and estimands have more policy-relevant interpretations compared to existing literature (20). By modelling the effect of intervening on mediators jointly we ensure that we are not violating model assumptions by adjusting for confounders of the mediator outcome pathway which are influenced by our exposure. We also carried out a sensitivity analysis to assess how resilient our estimates were to unmeasured confounding (21).

UKB is unrepresentative of the UK population and participants are less likely to live in socioeconomically deprived areas (30). Additionally, the methodology used to derive air pollution data excludes all participants living more than 400 km from Greater London (11). Population-wide studies are needed to establish the applicability of our findings to all regions of the UK.

All air pollution measures were based on a one-year estimate (2010-2011) at the participants’ baseline address which doesn’t consider changes of address since 2011. Mediator data were obtained at baseline assessment, which took place prior to measurement of air pollution. This is not ideal, as exposure should precede mediating factors to establish temporality. Therefore, this study relies on the assumption that air pollution exposure is static, but air pollution likely varies over time. Future research should use a more comprehensive model of air pollution exposure, including historical data about air pollution based on each participant’s address history.

### Conclusions

We find that most of the association between dementia and PM_2·5_, NO_2_ and NO_x_ cannot be attributed to indirect pathways through four potentially modifiable mediators. Having a cardiovascular health condition, seeking psychiatric treatment for anxiety or depression, physical inactivity and social isolation mediate a small proportion of this relationship. However, intervening on these mediators in individuals exposed to high levels of air pollution would only result in a ∼1% reduction in their dementia risk. Other unmeasured mediators may account for this relationship, or it may be due to a direct effect of the pollution itself. Therefore, public health measures directly aiming to reduce air pollution or protect the population from inhaling it should be prioritised to reduce dementia incidence.

## Data Availability

This research was conducted using the UK Biobank resource (application 123335). The data are not publicly available due to participant confidentiality and data protection agreements, but can be accessed by approved researchers through the UK Biobank application process (https://www.ukbiobank.ac.uk/)

## Contributors

All authors contributed to the conceptualisation of the study and methodology design. KT and MH led the data-curation. KT conducted the formal analysis and data visualisation. EH, EA and NM provided supervision. KT wrote the original draft and all other authors contributed to reviewing and editing of the manuscript. KT and MH accessed and verified the underlying data. All authors approved the final version of the manuscript.

## Acknowledgements

We would like to thank Professor Bianca De Stavola and Martin Danka for their guidance on our implementation of interventional effects for mediation analysis using g-formula. KT was funded by the Medical Research Council (Grant MR/W006774/1).

