## Supplementary Materials for "Mediation of the relationship between air pollution and dementia: A UK Biobank study"

### Supplementary material

Participants without mediator data

N = 11,002

Participants without demographic data

N = 16,953

Participants with mediator data

N = 432,935

Participants without air pollution data

N = 41,290

Participants with demographic data

N = 443,937

Participants with air pollution data

N = 460,890

Original UK Biobank sample

N = 502,180

Supplementary Figure 1. STROBE diagram

|  | **Quintile** | **OR** | **LL** | **UL** |
| --- | --- | --- | --- | --- |
| PM_2.5_ | 0 |  |  |  |
|  | 1 | 1.11 | 1.03 | 1.19 |
|  | 2 | 1.10 | 1.02 | 1.18 |
|  | 3 | 1.18 | 1.10 | 1.27 |
|  | 4 | 1.15 | 1.06 | 1.24 |
| NO_2_ | 0 |  |  |  |
|  | 1 | 1.09 | 1.02 | 1.17 |
|  | 2 | 1.16 | 1.08 | 1.25 |
|  | 3 | 1.09 | 1.01 | 1.18 |
|  | 4 | 1.08 | 1.00 | 1.18 |
| NO_x_ | 0 |  |  |  |
|  | 1 | 1.12 | 1.04 | 1.20 |
|  | 2 | 1.08 | 1.01 | 1.16 |
|  | 3 | 1.14 | 1.06 | 1.23 |
|  | 4 | 1.12 | 1.04 | 1.21 |
| PM_10_ | 0 |  |  |  |
|  | 1 | 1.10 | 1.03 | 1.18 |
|  | 2 | 1.14 | 1.06 | 1.22 |
|  | 3 | 1.08 | 1.00 | 1.16 |
|  | 4 | 1.03 | 0.96 | 1.11 |
| PM_2.5 Absorbance_ | 0 |  |  |  |
|  | 1 | 1.07 | 0.99 | 1.14 |
|  | 2 | 1.11 | 1.04 | 1.19 |
|  | 3 | 1.05 | 0.97 | 1.13 |
|  | 4 | 1.00 | 0.93 | 1.08 |
| PM_2.5-10_ | 0 |  |  |  |
|  | 1 | 1.03 | 0.96 | 1.10 |
|  | 2 | 1.01 | 0.94 | 1.08 |
|  | 3 | 1.00 | 0.93 | 1.08 |
|  | 4 | 0.97 | 0.91 | 1.05 |

Supplementary Table 1. Odds ratio of dementia incidence in association with air pollution quintile

| **Mediator** | **OR** | **LL** | **UL** |
| --- | --- | --- | --- |
| Cardiovascular condition | 1.71 | 1.62 | 1.80 |
| Psychiatric treatment | 1.47 | 1.38 | 1.57 |
| Insufficient exercise | 1.11 | 1.01 | 1.21 |
| Social isolation | 1.29 | 1.19 | 1.39 |

Supplementary Table 2. Odds ratio of dementia incidence in association with mediators.

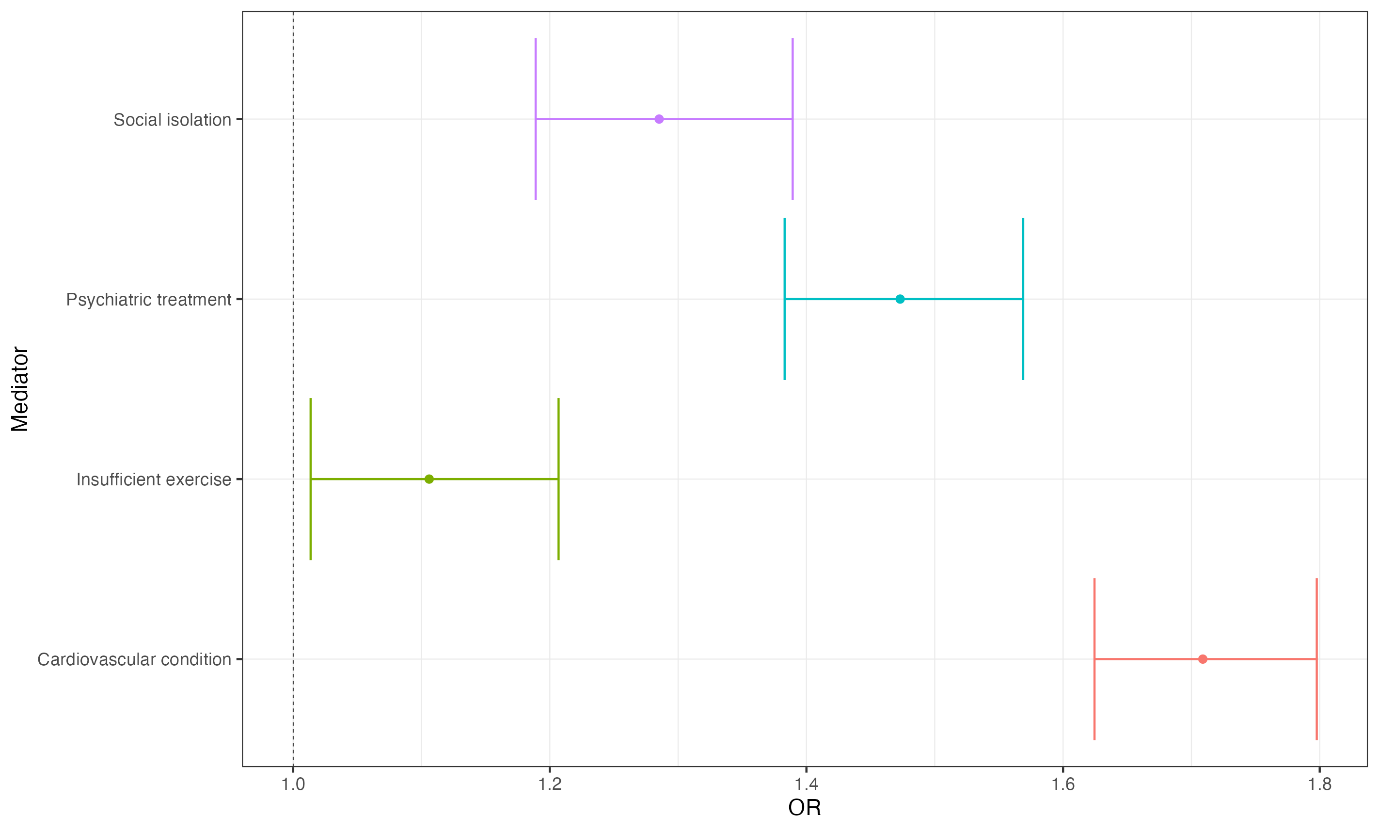

Supplementary Figure 2. Odds ratio of dementia incidence in association with mediators.

|  | | **Cardiovascular condition** | | | **Psychiatric treatment** | | | **Insufficient exercise** | | | **Social  isolation** | | |
| --- | --- | --- | --- | --- | --- | --- | --- | --- | --- | --- | --- | --- | --- |
| **Exposure** | **Quintile** | **OR** | **LL** | **UL** | **OR** | **LL** | **UL** | **OR** | **LL** | **UL** | **OR** | **LL** | **UL** |
| PM_2.5_ | 0 |  |  |  |  |  |  |  |  |  |  |  |  |
|  | 1 | 1.07 | 1.05 | 1.09 | 1.06 | 1.03 | 1.10 | 1.00 | 0.97 | 1.04 | 0.99 | 0.96 | 1.03 |
|  | 2 | 1.10 | 1.08 | 1.12 | 1.07 | 1.03 | 1.10 | 0.99 | 0.95 | 1.03 | 1.00 | 0.97 | 1.04 |
|  | 3 | 1.11 | 1.09 | 1.13 | 1.06 | 1.03 | 1.10 | 0.96 | 0.93 | 1.00 | 0.98 | 0.94 | 1.01 |
|  | 4 | 1.10 | 1.07 | 1.12 | 1.10 | 1.07 | 1.14 | 1.00 | 0.96 | 1.04 | 1.00 | 0.96 | 1.04 |
| NO_2_ | 0 |  |  |  |  |  |  |  |  |  |  |  |  |
|  | 1 | 1.09 | 1.07 | 1.11 | 1.04 | 1.01 | 1.08 | 0.96 | 0.92 | 0.99 | 0.95 | 0.91 | 0.98 |
|  | 2 | 1.13 | 1.11 | 1.16 | 1.07 | 1.04 | 1.10 | 0.98 | 0.95 | 1.02 | 0.91 | 0.88 | 0.94 |
|  | 3 | 1.10 | 1.08 | 1.12 | 1.09 | 1.06 | 1.13 | 0.98 | 0.95 | 1.02 | 0.90 | 0.87 | 0.93 |
|  | 4 | 1.02 | 1.00 | 1.04 | 1.11 | 1.07 | 1.15 | 0.98 | 0.94 | 1.02 | 0.99 | 0.95 | 1.03 |
| NO_x_ | 0 |  |  |  |  |  |  |  |  |  |  |  |  |
|  | 1 | 1.08 | 1.06 | 1.10 | 1.05 | 1.01 | 1.08 | 0.98 | 0.94 | 1.02 | 0.97 | 0.94 | 1.01 |
|  | 2 | 1.11 | 1.09 | 1.14 | 1.07 | 1.04 | 1.11 | 0.98 | 0.95 | 1.02 | 0.95 | 0.92 | 0.99 |
|  | 3 | 1.12 | 1.09 | 1.14 | 1.08 | 1.05 | 1.11 | 0.97 | 0.93 | 1.01 | 0.94 | 0.91 | 0.98 |
|  | 4 | 1.07 | 1.05 | 1.10 | 1.11 | 1.07 | 1.14 | 1.00 | 0.96 | 1.04 | 1.00 | 0.96 | 1.03 |
| PM_10_ | 0 |  |  |  |  |  |  |  |  |  |  |  |  |
|  | 1 | 1.05 | 1.03 | 1.07 | 1.03 | 1.00 | 1.06 | 1.03 | 1.00 | 1.07 | 0.99 | 0.96 | 1.03 |
|  | 2 | 1.04 | 1.02 | 1.06 | 1.04 | 1.01 | 1.07 | 0.99 | 0.95 | 1.02 | 0.98 | 0.95 | 1.02 |
|  | 3 | 1.03 | 1.01 | 1.05 | 1.05 | 1.02 | 1.09 | 1.05 | 1.01 | 1.08 | 1.02 | 0.98 | 1.05 |
|  | 4 | 1.04 | 1.02 | 1.06 | 1.05 | 1.02 | 1.08 | 1.03 | 0.99 | 1.07 | 1.04 | 1.00 | 1.07 |
| PM_2.5 Absorbance_ | 0 |  |  |  |  |  |  |  |  |  |  |  |  |
|  | 1 | 1.10 | 1.08 | 1.12 | 1.01 | 0.98 | 1.04 | 0.96 | 0.93 | 1.00 | 0.88 | 0.85 | 0.91 |
|  | 2 | 1.09 | 1.07 | 1.12 | 1.04 | 1.01 | 1.07 | 0.96 | 0.92 | 0.99 | 0.87 | 0.84 | 0.90 |
|  | 3 | 1.07 | 1.05 | 1.10 | 1.05 | 1.02 | 1.09 | 0.98 | 0.95 | 1.02 | 0.89 | 0.86 | 0.93 |
|  | 4 | 0.99 | 0.97 | 1.01 | 1.08 | 1.05 | 1.12 | 1.00 | 0.96 | 1.04 | 0.97 | 0.93 | 1.00 |

Supplementary Table 3. Odds ratio of mediators in association with air pollution quintile.

| **Exposure** | **Quintile** | **Effect** | **Estimate** | **95% CI** | |
| --- | --- | --- | --- | --- | --- |
| PM_2.5_ | 1 | Rpnde | 1.10 | 1.02 | 1.17 |
|  |  | Rpnie | 1.01 | 1.01 | 1.01 |
|  |  | Rte | 1.11 | 1.03 | 1.18 |
|  | 2 | Rpnde | 1.08 | 1.02 | 1.16 |
|  |  | Rpnie | 1.01 | 1.01 | 1.02 |
|  |  | Rte | 1.10 | 1.03 | 1.18 |
|  | 3 | Rpnde | 1.16 | 1.08 | 1.25 |
|  |  | Rpnie | 1.01 | 1.01 | 1.02 |
|  |  | Rte | 1.18 | 1.10 | 1.27 |
|  | 4 | Rpnde | 1.13 | 1.04 | 1.21 |
|  |  | Rpnie | 1.01 | 1.01 | 1.02 |
|  |  | Rte | 1.14 | 1.06 | 1.23 |
| NO_2_ | 1 | Rpnde | 1.08 | 1.01 | 1.16 |
|  |  | Rpnie | 1.01 | 1.01 | 1.01 |
|  |  | Rte | 1.09 | 1.02 | 1.17 |
|  | 2 | Rpnde | 1.14 | 1.06 | 1.22 |
|  |  | Rpnie | 1.02 | 1.01 | 1.02 |
|  |  | Rte | 1.16 | 1.08 | 1.23 |
|  | 3 | Rpnde | 1.08 | 1.01 | 1.16 |
|  |  | Rpnie | 1.01 | 1.01 | 1.02 |
|  |  | Rte | 1.09 | 1.02 | 1.17 |
|  | 4 | Rpnde | 1.07 | 0.99 | 1.16 |
|  |  | Rpnie | 1.01 | 1.00 | 1.01 |
|  |  | Rte | 1.08 | 0.99 | 1.16 |
| NO_x_ | 1 | Rpnde | 1.10 | 1.03 | 1.19 |
|  |  | Rpnie | 1.01 | 1.01 | 1.01 |
|  |  | Rte | 1.11 | 1.04 | 1.20 |
|  | 2 | Rpnde | 1.07 | 0.99 | 1.14 |
|  |  | Rpnie | 1.01 | 1.01 | 1.02 |
|  |  | Rte | 1.08 | 1.01 | 1.15 |
|  | 3 | Rpnde | 1.12 | 1.04 | 1.20 |
|  |  | Rpnie | 1.01 | 1.01 | 1.02 |
|  |  | Rte | 1.13 | 1.05 | 1.22 |
|  | 4 | Rpnde | 1.10 | 1.02 | 1.19 |
|  |  | Rpnie | 1.01 | 1.01 | 1.01 |
|  |  | Rte | 1.11 | 1.03 | 1.20 |

Supplementary Table 4. Mediation analysis decomposed effects. Rpnde: Randomised pure natural direct effect. Rpnie: Randomised pure natural indirect effect. Rte: Randomised total effect.

| **Exposure** | **Quintile** | **Effect** | **estRR** | **lowerRR** | **upperRR** | **Evalue.estRR** | **Evalue.lowerRR** |
| --- | --- | --- | --- | --- | --- | --- | --- |
| PM_2.5_ | 1 | Rpnde | 1.10 | 1.02 | 1.17 | 1.42 | 1.18 |
|  |  | Rpnie | 1.01 | 1.01 | 1.01 | 1.11 | 1.09 |
|  |  | Rte | 1.11 | 1.03 | 1.18 | 1.45 | 1.22 |
|  | 2 | Rpnde | 1.08 | 1.02 | 1.16 | 1.39 | 1.14 |
|  |  | Rpnie | 1.01 | 1.01 | 1.02 | 1.13 | 1.11 |
|  |  | Rte | 1.10 | 1.03 | 1.18 | 1.43 | 1.21 |
|  | 3 | Rpnde | 1.16 | 1.08 | 1.25 | 1.59 | 1.37 |
|  |  | Rpnie | 1.01 | 1.01 | 1.02 | 1.13 | 1.11 |
|  |  | Rte | 1.18 | 1.10 | 1.27 | 1.63 | 1.42 |
|  | 4 | Rpnde | 1.13 | 1.04 | 1.21 | 1.50 | 1.26 |
|  |  | Rpnie | 1.01 | 1.01 | 1.02 | 1.13 | 1.11 |
|  |  | Rte | 1.14 | 1.06 | 1.23 | 1.54 | 1.31 |
| NO_2_ | 1 | Rpnde | 1.08 | 1.01 | 1.16 | 1.38 | 1.11 |
|  |  | Rpnie | 1.01 | 1.01 | 1.01 | 1.11 | 1.09 |
|  |  | Rte | 1.09 | 1.02 | 1.17 | 1.41 | 1.16 |
|  | 2 | Rpnde | 1.14 | 1.06 | 1.22 | 1.54 | 1.33 |
|  |  | Rpnie | 1.02 | 1.01 | 1.02 | 1.14 | 1.12 |
|  |  | Rte | 1.16 | 1.08 | 1.23 | 1.58 | 1.38 |
|  | 3 | Rpnde | 1.08 | 1.01 | 1.16 | 1.37 | 1.10 |
|  |  | Rpnie | 1.01 | 1.01 | 1.02 | 1.12 | 1.10 |
|  |  | Rte | 1.09 | 1.02 | 1.17 | 1.40 | 1.17 |
|  | 4 | Rpnde | 1.07 | 0.99 | 1.16 | 1.35 | 1.00 |
|  |  | Rpnie | 1.01 | 1.00 | 1.01 | 1.08 | 1.05 |
|  |  | Rte | 1.08 | 0.99 | 1.16 | 1.37 | 1.00 |
| NO_x_ | 1 | Rpnde | 1.10 | 1.03 | 1.19 | 1.44 | 1.19 |
|  |  | Rpnie | 1.01 | 1.01 | 1.01 | 1.11 | 1.08 |
|  |  | Rte | 1.11 | 1.04 | 1.20 | 1.47 | 1.23 |
|  | 2 | Rpnde | 1.07 | 0.99 | 1.14 | 1.33 | 1.00 |
|  |  | Rpnie | 1.01 | 1.01 | 1.02 | 1.13 | 1.11 |
|  |  | Rte | 1.08 | 1.01 | 1.15 | 1.38 | 1.08 |
|  | 3 | Rpnde | 1.12 | 1.04 | 1.20 | 1.48 | 1.25 |
|  |  | Rpnie | 1.01 | 1.01 | 1.02 | 1.13 | 1.11 |
|  |  | Rte | 1.13 | 1.05 | 1.22 | 1.52 | 1.30 |
|  | 4 | Rpnde | 1.10 | 1.02 | 1.19 | 1.44 | 1.16 |
|  |  | Rpnie | 1.01 | 1.01 | 1.01 | 1.11 | 1.10 |
|  |  | Rte | 1.11 | 1.03 | 1.20 | 1.47 | 1.20 |

Supplementary Table 5. Sensitivity analysis for unmeasured confounding in mediation analysis.
